# The post-pandemic hospital and mortality burden of COVID-19 compared with influenza: A national cohort study in Denmark, May 2022 to June 2024

**DOI:** 10.1101/2024.09.26.24314428

**Authors:** Peter Bager, Ingrid Bech Svalgaard, Frederikke Kristensen Lomholt, Hanne-Dorthe Emborg, Lasse Engbo Christiansen, Bolette Soborg, Anders Hviid, Lasse Skafte Vestergaard

**Affiliations:** Department of Infectious Disease Epidemiology and Prevention, Statens Serum Institut, Copenhagen, Denmark; Department of Epidemiology Research, Statens Serum Institut, Copenhagen, Denmark; Pharmacovigilance Research Center, Department of Drug Design and Pharmacology, University of Copenhagen, Copenhagen, Denmark

**Keywords:** COVID-19, SARS-CoV-2, influenza, burden, admissions, mortality, pandemic

## Abstract

**Background:** In the post-pandemic period, COVID-19 continues to cause significant numbers of hospitalisations and deaths. We describe this burden and compare it to the burden of influenza in the first two post-pandemic years in Denmark.

**Methods:** A cohort study including residents in Denmark from May 16, 2022, to June 7, 2024. Data were obtained from national registries, including information on Polymerase chain reaction (PCR) test-positive COVID-19 and influenza admissions, mortality within 30 days of admission, sex, age, COVID-19 and influenza vaccination, comorbidity, and living in long-term care facility for elderly. Negative binomial regression was used to estimate adjusted incidence rate ratios (aIRRs) to compare rates of admissions between COVID-19 and influenza. To assess severity of COVID-19 among hospitalized patients, we used Cox proportional hazard models to estimate adjusted hazard ratios (aHR) of 30-day mortality between COVID-19 and influenza.

**Results:** Among 5,899,170 individuals, admissions with COVID-19 (n=24,687) were more frequent than admissions with influenza (n=8,682; aIRR 2.01, 95%CI 1.37-2.95), in particular during the first year (p=0.01), in the summer (p<0.001) and among people above 65 years of age (p<0.001). The number of deaths were also higher (COVID-19, n=2,393; influenza, n=522). Among patients, the risk of mortality of COVID-19 was higher than influenza in the 12-30 days following admission (0-11 days, aHR 1.08, 95%CI 0.94-1.25; 12-30 days, aHR 1.50, 95%CI 1.21-1.84), in particular among non-vaccinated for both COVID-19 and influenza (aHR 1.81, 95%CI 1.25-2.62), while it was similar to influenza among patients without comorbidities (aHR 1.07, 95%CI 0.63-1.80).

**Conclusion:** COVID-19 represented a greater disease burden than influenza, with more hospitalisations and deaths, and more severe disease primarily among non-vaccinated and comorbid patients. These results highlight the continued need for attention and public health efforts to mitigate the impact of SARS-CoV-2.

## INTRODUCTION

As the COVID-19 pandemic subsides, infections with SARS-CoV-2 persist, resulting in a substantial seasonal hospital and mortality burden, but moving towards resembling that of seasonal influenza. With decades of established data on influenza epidemiology, burden, and management, it is an ideal benchmark against which to assess COVID-19’s evolving impact, in turn informing public health strategies, resource allocation, and preparedness efforts.^1^

Only few studies have compared the post-pandemic disease burden of COVID-19 with influenza, all suggesting a higher disease severity of COVID-19, as measured by 30-day mortality among hospitalized patients. However, generalizability was limited, because the studies included mostly elderly male patients from the United States Veterans Affairs health care system.^2,3^

In this study, we utilized data for the entire general population of Denmark to compare COVID-19 versus influenza from 2022 to 2024, to assess both the number of admissions and deaths, and to compare disease severity.

## METHODS

### Study design and participants

This nationwide observational cohort study included all residents of Denmark in the COVID-19 post-pandemic period, defined as from May 16, 2022 to June 7, 2024, thus applying the definition of influenza season months of the European and US centers’ for disease control and prevention.^4,5^ Participants were considered hospitalized with COVID-19 or influenza if they had a positive PCR test from 14 days before and up to two days after their hospital admission date. The date of admission was used to calculate 30-day mortality.

### Ethics

The study was conducted as part of Statens Serum Institut’s governmental duties as National Public Health Agency. According to Danish law, ethical approval or individual consent is exempt for anonymised aggregated register-based studies, such as the present. Due to the nature of this research, there was no direct involvement of patients or members of the public in the design or reporting of this study. Therefore, no approval from an ethics committee is required. The study is fully compliant with all legal and ethical requirements and there are no further processes available regarding such studies.

### Data sources

The study was based on Denmark’s national COVID-19 and influenza surveillance system and population-based registers with individual-level data that are updated and linked daily using the unique civil registration number given to all residents.^6^ Details of all COVID-19 and influenza vaccinations administered in the country were obtained from the Danish National Vaccination Register.^7^ Hospital admission and discharge dates, diagnoses and referrals were obtained from the National Patient Registry.^8^ Data on comorbidities based on the International Classification of Diseases, tenth revision, diagnosis codes (haematological disease, cancers, neurological diseases, kidney diseases, cardiovascular diseases, respiratory diseases, and immunological conditions) were obtained from the National Patient Registry, and data on additional comorbidities (asthma, dementia, type 1 diabetes, type 2 diabetes, chronic obstructive pulmonary disease, rheumatoid arthritis, osteoporosis, schizophrenia) were obtained from the Register of Selected Chronic Diseases and Severe Mental Disorders.^9^ Data on living in a long-term care facility (LTCF) for older people (the majority being older than 65 years) were obtained from the LTCF address database.^10^ Death dates, where applicable, and residency data were obtained from the Civil Registration System^6^ along with information on sex, age, address history, emigration, and disappearance from national registers. In adherence to Sex and Gender Equity in Research (SAGER) guidelines for reporting,^11^ we derived the sex variable from the civil registration numbers assigned at birth or time of entry to Denmark. People with an odd numerical value are categorised as male at birth, while those with an equal numerical value are categorized as female. This method aligns with SAGER guidelines, providing a standardized approach to sex assignment in our analysis. Details of all SARS-CoV-2 and influenza PCR tests conducted during the study period were obtained from the Danish Microbiology Database^12^ and originated from one of three sources: Routine clinical samples collected on indication at hospitals and primary care clinics country-wide,^13^ clinical samples collected on indication in selected primary care clinics as part of the national sentinel surveillance system for influenza-like illness (ILI) based on ECDC and WHO guidelines,^14^ and a self-sampling virus monitoring system, known as Virus monitoring in Denmark, implemented since May 2022.^15,16^ Clinical samples collected on indication at hospitals and primary care clinics were analyzed by one of the country’s ten Departments of Microbiology, and Virus monitoring samples were analyzed by Statens Serum Institut.^13^

### Test strategy, mass-vaccinations and circulating virus strains

February 1, 2022, marked the official date for removal of all COVID-19 restrictions in Denmark, with a continued downscaling of a world-record high test-level (4 million weekly PCR and antigen tests) to a lower level on March 7 and onwards, with weekly SARS-CoV-2 PCR tests ranging from 35-72.000 in 2022 to 3.000-11.000 in 2023 and 2024); the parallel weekly influenza PCR tests ranged from 4000-11.000.^17,18^ Since March 10, 2022, the health authorities recommended PCR-tests only for persons with symptoms and at risk of a severe disease course of COVID-19, and this recommendation was identical for influenza.^19^

Mass-vaccination and circulating virus strains just before the end of the pandemic are briefly described in the supplement, text B. After the pandemic, in the winter season 2022-23, mass-vaccination targeting individuals aged 65 years and above was rolled out on October 1, 2022. More than 95% of adults above 65 years received COVID-19 booster doses targeting both the ancestral strain and either omicron subvariant BA.1 or BA.4/5.^20,21^ To protect against influenza, more than 81% of adults above 65 years received the quadrivalent influenza vaccine targeting the A(H1N1), A(H3N2), B-Victoria, and B-Yamagata strains.^20^ During the winter season, infections with recombinant XBB subvariants of omicron became dominant over other subvariants including BA.1 and BA.4/5, while influenza A(H1N1,) (42.2%), A(H3N2) (19.8%) and B (38.0%) infections co-circulated.^22^ In the following winter season, 2023-24, mass-vaccination again started on October 1, 2023, and adults above 65 years of age were offered booster doses targeting the XBB.1.5 subvariant, now with *simultaneous* administration of the quadrivalent influenza vaccine; more than 78% received the two vaccines.^20^ During that winter season, infections with the hyper-mutated new omicron subvariant BA.2.86 began to dominate,^16,22^ while influenza A(H1N1) infections dominated (72.1%), followed by A(H3N2) (27.8%).

### Statistical analysis

*Nationwide disease burden*: We used negative binomial regression to estimate sex- and age-adjusted incidence rate ratios (aIRRs) to compare rates of admissions between COVID-19 and influenza among all individuals living in Denmark, as described in detail in the supplement (page xx). *Comparative 30-day mortality after hospitalisation:* We analysed disease severity (approximated as 30-day mortality) among individuals living in Denmark who were hospitalized with either COVID-19 or influenza, with the positive test taken between December and May of 2022-2023 and 2023-2024. We thereby excluded individuals admitted in the summer and fall (June to November), for brevity “summer season”. This was done due to too small influenza mortality numbers for meaningful comparison with COVID-19. Furthermore, individuals with missing information of region of residence were excluded due to small numbers. The cohort was followed for 30 days or until death, emigration, disappearance from national registers or the end of study period, whichever occurred first. Baseline characteristics between patients hospitalized with COVID-19 and influenza were compared using standardized mean differences (SMD), with a SMD < 0.1 indicating good balance.^23^ An overview of baseline variables is provided in Table S1. Differences in baseline characteristics between the two groups were adjusted for using inverse probability weighting. A propensity score (PS), calculated using logistic regression to estimate the probability of being assigned to the COVID-19 group, was applied to balance the groups. Weights were constructed as 1 for COVID-19 group and PS/(1-PS) for the influenza group. Both unweighted and weighted Kaplan-Meier curves were constructed. Additionally, the risk difference (RD) was estimated using the cumulative incidences at 30 days since admission produced by the weighted Kaplan-Meier estimator with corresponding 95% confidence intervals calculated by the delta method. A weighted Cox proportional hazards model, with time since admission as the underlying time scale, was used to estimate the hazard of death between COVID-19 and influenza groups, with results reported as adjusted hazard ratios (aHRs) with 95% confidence intervals (CI). The proportional hazards assumption was assessed using the scaled Schoenfeld residuals, both graphically and by testing for independence between residuals and time. If the assumption was violated, appropriate time points were determined based on these, and step-wise models were fitted. We conducted stratified analyses based on the following variables: Sex, age (above or below 65 years of age), comorbidity (any; none) and vaccination status within 180 days of the test-date of the admission (vaccinated against both diseases; vaccinated against the disease the individual tested positive for; not vaccinated). A sensitivity analyses was conducted to investigate immortal time bias by setting the start of follow-up as the date of the PCR test for individuals who tested positive after admittance to the hospital in the analysis of disease severity. All analyses were carried out in R version 4.1.1.^24^ The packages MASS (v7.3-54), performance (v0.12.0), and survival (v3.2-11) were used for modelling and ggplot2 (v3.5.1) and forestploter (v1.1.1) for visualizations.^25-29^

## RESULTS

The study cohort consisted of 5,899,170 individuals living in Denmark during the first two post-pandemic years, from May 16, 2022, to June 7, 2024. During this period, 618,470 (10.5%) individuals were tested for COVID-19 and/or influenza, of whom 334,335 (54.1%) tested positive. Among those who tested positive, 32,054 (9.6%) were defined as admitted to the hospital with COVID-19 (n=24,687) or influenza (n=8,682), constituting 33,369 admissions (Figure S1).

32,212 (96.5%) of the admitted patients had their samples collected at hospitals or primary care clinics, as the PCR-test were performed by Departments of Microbiology (see Methods). The majority of patients were tested both for COVID-19 and influenza on the same date in connection with the admission, i.e. 82.1% of COVID-19 patients and 97.8% for influenza patients (see Table S2). Among influenza patients, 7,623 had influenza A (87.8%) and 1,059 influenza B (12.4%).

The mean length of hospital stays was slightly longer for patients with COVID-19 (4.71 days) compared to patients with influenza (4.08 days). While the median length of stay was 3 days for both patient groups, COVID-19 patients exhibited slightly longer stays at the upper end of the distribution, with 95% of patients staying ≤ 13 days compared to ≤ 12 days for patients with influenza. (Table S3).

### Nationwide disease burden of COVID-19 vs. influenza admissions and deaths

Overall, the absolute number of COVID-19 admissions and deaths was higher when compared to influenza (24,687 vs. 8,682 admissions, and 2,393 vs. 522 deaths). This was the case during most months of the two years (except for February 2023 and February 2024, see Figure 1). The difference was more pronounced in the first year, during the summer season, among males, and among patients older than 65 years (Table 1). These results were consistent with the sex- and age-adjusted IRRs based on 11.9 million person-years of follow-up. The admission rate for COVID-19 was higher than for influenza (aIRR 2.01, 95%CI 1.37-2.95), with the difference more pronounced in the first year compared to the second (aIRR 2.63, 95%CI 1.72-4.01, p=0.01), during the summer season (aIRR>14, p<0.001), and among people older than 65 years (aIRR 4.30, 95%CI 3.28-5.63, p<0.001), however, there were no significant difference between males and females (p=0.71) (see Table S4).

**Table 1.** Characteristics and adjusted incidence rate ratios of admissions (n=33,369) and mortality (n=2,915) comparing patients with COVID-19 and influenza among the Danish population (n=5.9 mio., 11.9 million person-years of follow-up), May 2022 to June 2024.

|  | **Admission**<br>(positive virus PCR test up to 14 days before or 2 days<br>after admission date) |  | **Mortality**<br>(30-day mortality after admission date) |  |
| --- | --- | --- | --- | --- |
|  | COVID_19 | Influenza | COVID-19 | Influenza |
|  | n (%) | n (%) | n (%) | n (%) |
| **Total** | 24687 (100%) | 8682 (100%) | 2393 (100%) | 522 (100%) |
| **Year\*** |  |  |  |  |
| 2022/2023 | 16181 (65.5%) | 3973 (45.8%) | 1510 (63.1%) | 212 (40.6%) |
| 2023/2024 | 8506 (34.5%) | 4709 (54.2%) | 883 (36.9%) | 310 (59.4%) |
| **Season** |  |  |  |  |
| Summer-fall 2022 | 9210 (37.3%) | 173 (2.0%) | 796 (33.3%) | 10 (1.9%) |
| Winter-spring<br>2022/2023 | 7239 (29.3%) | 3820 (44.0%) | 736 (30.8%) | 202 (38.7%) |
| Summer-fall 2023 | 4271 (17.3%) | 213 (2.5%) | 376 (15.7%) | 13 (2.5%) |
| Winter-spring<br>2023/2024 | 3967 (16.1%) | 4476 (51.6%) | 485 (20.3%) | 297 (56.9%) |
| **Sex** |  |  |  |  |
| Female | 11446 (46.4%) | 4408 (50.8%) | 1017 (42.5%) | 262 (50.2%) |
| Male | 13241 (53.6%) | 4274 (49.2%) | 1376 (57.5%) | 260 (49.8%) |
| **Age** |  |  |  |  |
| 0-39 | 1989 (8.1%) | 2027 (23.3%) | 11 (0.5%) | 14 (2.7%) |
| 40-65 | 4128 (16.7%) | 2152 (24.8%) | 168 (7.0%) | 62 (11.9%) |
| 65+ | 18570 (75.2%) | 4503 (51.9%) | 2214 (92.5%) | 446 (85.4%) |

Summer-fall, summer season; winter-spring, winter season. \* 2022/2023: May 16 2022 – May 16 2023, 2023-2024: May 16 2023 – study end

**Figure 1.**
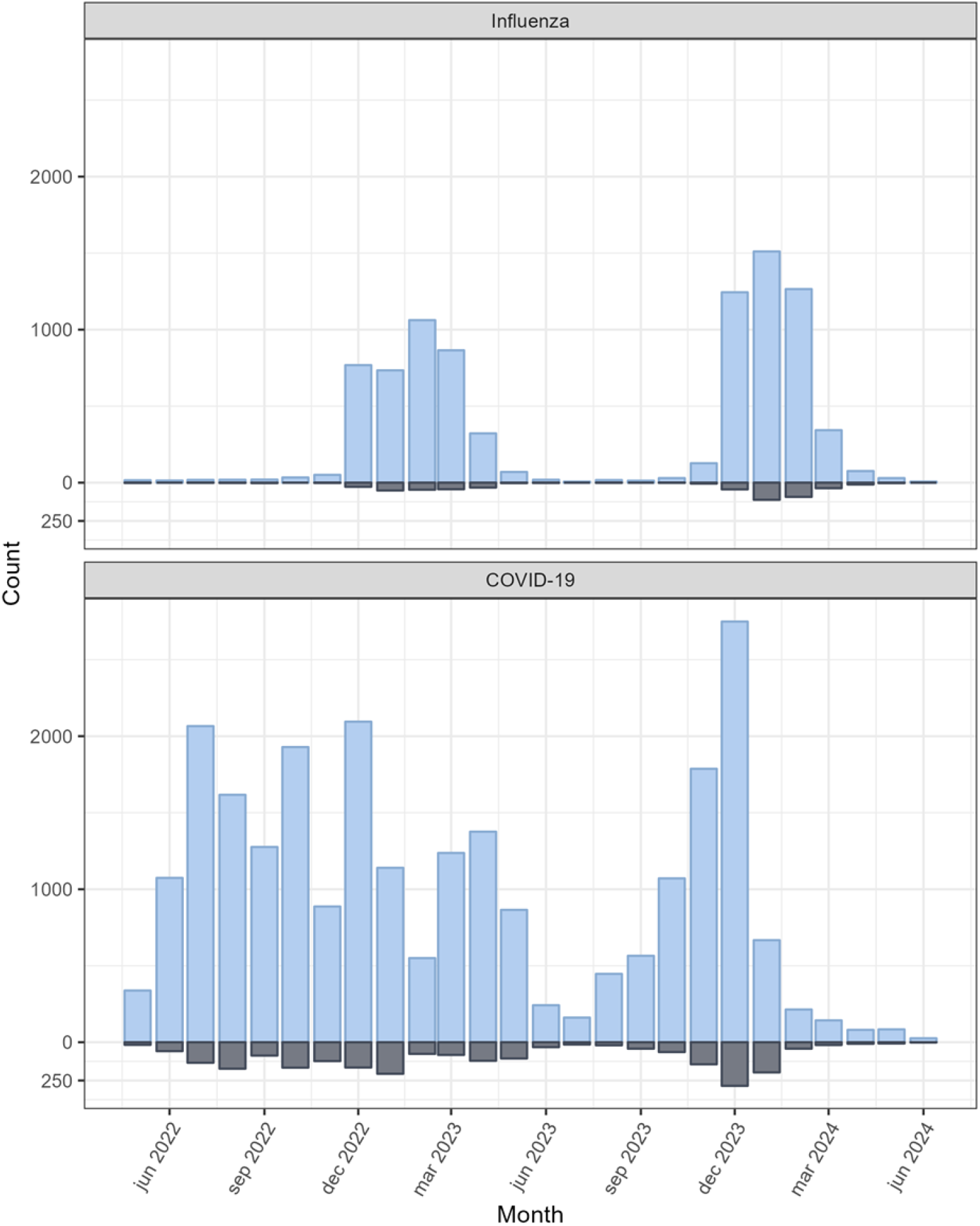
Number of COVID-19 and influenza admissions (blue) and 30-day mortalities (grey) by month, May 2022 to June 2024, Denmark.

### Disease severity of COVID-19 vs. influenza

We restricted the study population to individuals admitted in the winter and spring (December to May), for brevity “winter season”, (COVID-19, n=11,515; influenza, n=8,298) and excluded individuals admitted in the summer season (COVID-19, n=13,172; influenza, n=384). Characteristics of included and excluded patients are shown in Table S5. The most notable difference were more vaccinations among the included vs. excluded patients, which was due to the definition of vaccination *within* six months resulting in the winter season patients more recently being part of the mass-vaccination from October into January, as opposed to the summer season patients.

Figure 2 shows Kaplan Meier 30-day survival curves for COVID-19 patients and with influenza patients both unweighted and weighted to the distributions of the baseline characteristics of the COVID-19 patients. The unweighted analysis showed lower survival among COVID-19 patients throughout the 30 days following admission (day 30: 89.2% vs. 94.0% for influenza). However, in the weighted analysis, survival was more similar during the first 10-12 days, and then survival of COVID-19 patients continued to decline from day 12 to 30, whereas it started to level off for influenza patients, resulting in non-overlapping confidence intervals after 23 days (day 30: 89.2% vs. 91.1% for influenza). There was a higher risk of death within 30 days of admission among patients with COVID-19 compared to those with influenza (RD 1.86 95% CI 0.79, 2.93) after adjusting for differences in patient characteristic between the groups. This translates to an estimated 19 (95% CI 8-29) additional deaths per 1000 patients hospitalized with covid-19 compared to those hospitalized with influenza.

**Figure 2.**
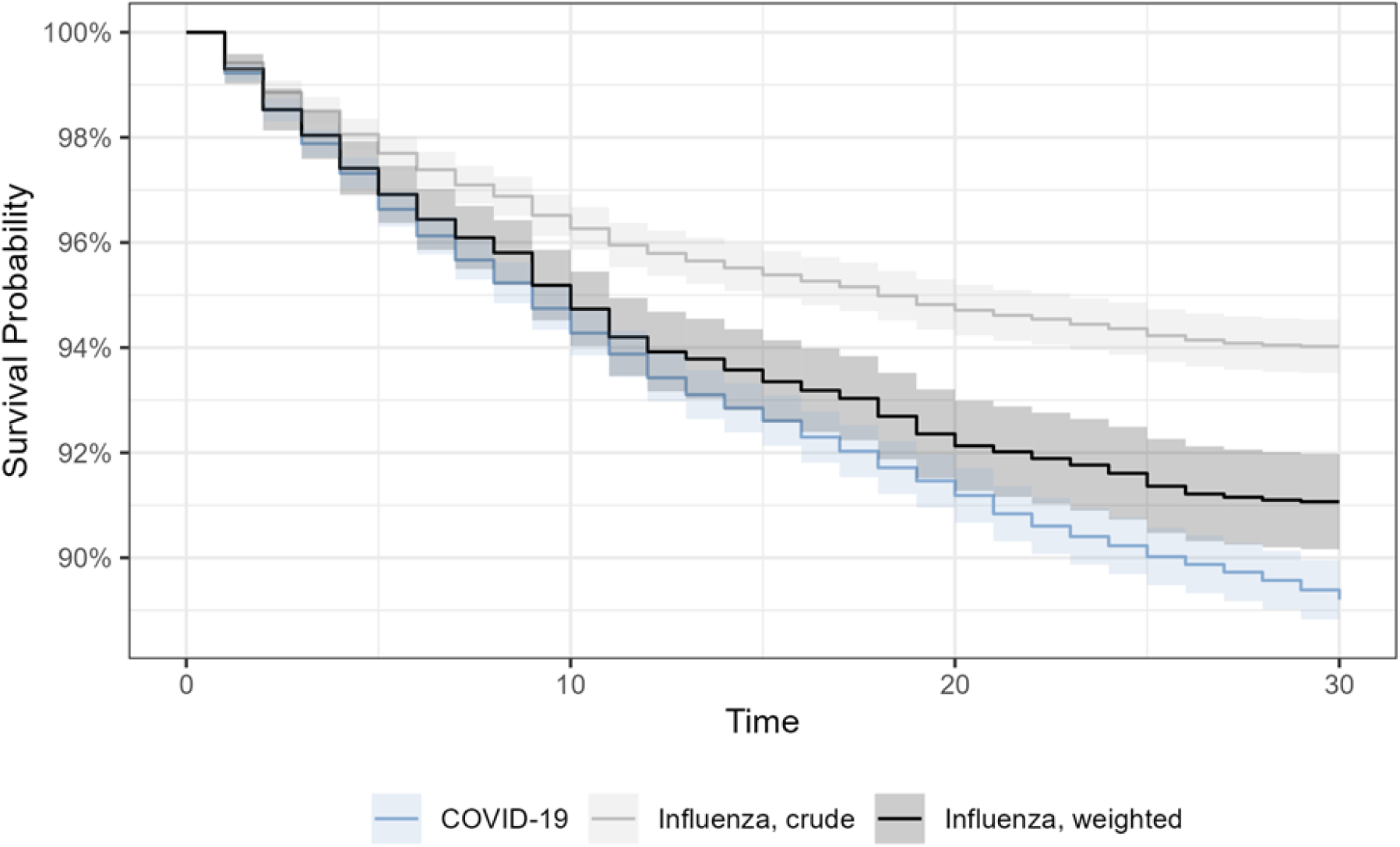
Kaplan-Meier survival curve for 11,515 COVID-19 patients and 8,298 influenza patients, both crude and weighted. The patient population is restricted to those with infections during the winter season 2022-2024, Denmark.

The adjusted HR for 30-day mortality was slightly higher overall for COVID-19 compared to influenza (aHR 1.22, 95% CI 1.08-1.37) but concealed a time-dependent effect (i.e. detected as proportional hazards assumption violation). Specifically, the increased risk was observed 12-30 days after admission (0-11 days, aHR 1.08, 95%CI 0.94-1.25; 12-30 days, aHR 1.50, 95%CI 1.21-1.84) (Figure 3). In analyses stratified by sex, age, comorbidity and vaccination, the increased risk of mortality 12-30 days after admission was significant for patients above 65 years of age but not below (aHR 1.49, 95%CI 1.19-1.86 vs. aHR 1.48, 95%CI 0.80-2.73), primarily for patients with comorbidities compared to those without (aHR 1.61, 95%CI 1.28-2.03 vs. aHR 1.07, 95%CI 0.63-1.80), and slightly higher for patients neither vaccinated against COVID-19 nor influenza (aHR 1.81, 95%CI 1.25-2.62 vs. aHR 1.37, 95%CI 1.05-1.79, for COVID-19 and influenza vaccinated, and aHR 1.37, 95%CI 1.05-1.79, for COVID19 or influenza vaccinated). Stratification on sex showed similar risk of mortality (male, aHR 1.61, 95%CI 1.21-2.15; female, aHR 1.40, 95%CI 1.03-1.90). (Figure 3).

**Figure 3.**
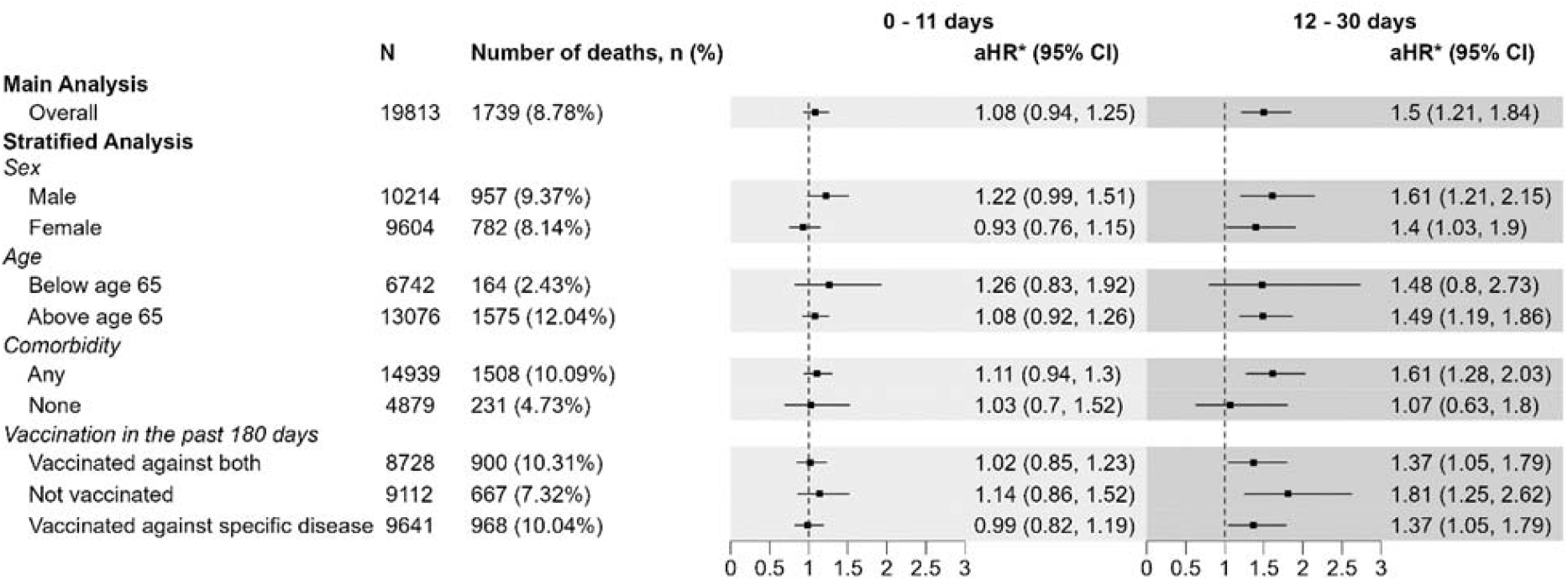
Risk of mortality within 30 days of admission for 11,515 COVID-19 compared to 8,298 influenza patients stratified by time since admission (0-11 days, 12-30 days) and further stratified with regards to baseline characteristics. The patient population is restricted to those with infections during the winter and spring, 2022-2024, Denmark. Adjusted hazard ratios (aHR) and 95% confidence intervals (CI). *differences in baseline characteristics between the groups were adjusted for using inverse probability weighting. Logistic regression was used to calculate a propensity score (probability of being assigned to the COVID-19 group).

### Sensitivity analyses

In a sensitivity analysis investigating immortal time bias, results for disease severity were similar to the main finding (0-11 days, aHR 1.08, 95%CI 0.93-1.25; 12-30 days, aHR 1.50, 95%CI 1.20-1.84). Sensitivity analyses of IRR and HR using influenza A and B as comparison groups was not meaningful due to small numbers for influenza B (e.g. only 26 died within 30 days of admission).

## DISCUSSION

We found that COVID-19 during the first two years after the pandemic like seasonal influenza leads to a considerable winter-burden of admissions and mortality in Denmark. However, the burden has been larger than for influenza, in particular in the first post-pandemic year, and especially in the summer when influenza is rare, and among elderly. In the winter season, admission numbers were more similar to influenza, but there were more COVID-19 deaths, and the COVID-19 admissions were more serious, primarily among non-vaccinated and comorbid patients.

Our finding of a more than 2-fold larger burden of hospital admissions for COVID-19 than influenza during the first two post-pandemic winter seasons, adjusted for age and sex-differences, corresponds well to the crude ratio based on surveillance data from e.g. the Centers for Disease Control and Prevention, USA.[17, 51, 52][53]. A key question is whether the COVID-19 burden will become smaller and smaller in the post-pandemic years. With only two years of data, however, this is yet difficult to establish. In the second year, we observed a decrease in the number of admissions and deaths with COVID-19, although still higher than for influenza. Crude surveillance data from other countries, e.g. USA, Ireland, and New Zealand, also show a decrease in COVID-19 admissions (May 2022 to June 2024),^30-32^ while wastewater signals of infection levels (copies of DNA per person per day) show no consistent decrease and even suggest near pandemic-high levels. ^33-36^ One straightforward explanation for the decrease in admissions is that individuals at higher risk of a severe clinical outcomes of COVID-19 are offered protection through the annual COVID-19 vaccination program. However, we are inclined to mention two observations not consistent with a decrease in admissions in the second year: Firstly, COVID-19 vaccination coverage in elderly in Denmark decreased from 95% in the first year, to 78% in the second year, where the speed of roll-out was also slower,^37^ and secondly, we observed a slightly lower vaccine effectiveness against admission with the new SARS-CoV-2 variants emerging in the second year (the hypermutated BA.2.86 and it’s subvariant JN.1) relative to previous variants that year. ^16^ On the other hand, the two observations should be viewed on the background that the vaccination coverage in Denmark was the highest in Europe, and protected throughout the winter.^37^

With regards to our observations on disease severity, previous studies have also suggested that COVID-19 admissions are more severe than influenza - also using 30-day mortality as indicator of severe disease courses. Thus, among elderly male patients in the US veterans’ cohort, two studies reported 61% higher 30-day mortality in the first post-pandemic year (HR 1.61, 95% CI 1.29-2.02, n=11,399), and 35% higher in the second year (HR 1.35, 95% CI 1.10-1.66, n=11,272), when compared to influenza admissions.^2,3^ In our larger general patient population from Denmark (n=33,369), the differences were more modest, with no difference in risk of mortality from day 1 to 11 after admission, but from day 12 to 30 a 50% higher risk of mortality among patients with COVID-19 compared to influenza. On the other hand, it has been argued that COVID-19 and influenza has marginal differences in disease severity based on crude fatality rates ranging only from 0.05 to 0.10 (i.e. comparable to 0.94-1=0.06 in our Figure 3).^38^ We observed approximately 19 more deaths per 1,000 admissions with COVID-19 relative to influenza. A potential reason for the higher mortality could be that COVID-19, compared to influenza, is characterized by more persistent respiratory symptoms, and more systemic effects, e.g. gastrointestinal symptoms and different body dysfunctions (olfactory, cardiac, hepatobiliary, renal) likely as a result of viral or indirect injury to the tissue cells, although the pathophysiological mechanisms are not fully understood.^39,40^

In stratified analyses, we found that the higher risk of mortality at day 12-30 for COVID-19 patients than influenza patients was even greater among patients who were neither vaccinated against COVID-19 nor influenza. This finding not only stresses the important role of the seasonal booster vaccination program to elderly above 65 years, but also the role of inadequate COVID-19 immunity in younger age-groups, constituting more than half of this un-vaccinated group. Another important observation was that among non-comorbid patients, e.g. patients without cardiac disease, chronic obstructive pulmonary disease, diabetes, the risk of mortality was similar for COVID-19 and influenza, while COVID-19 patients with comorbidities still had more serious admissions than those with influenza.

We took advantage of clinically collected PCR test samples in order to similarly define COVID-19 and influenza admission and deaths. This was done to circumvent bias from the alternative of using diagnoses and causes of death registrations that are not sufficiently uniform and validated for this comparison. A major advantage of the study was that test-recommendations for COVID-19 and influenza included similar clinical indications in the two post-pandemic years, i.e. test of persons with symptoms and at risk of severe COVID-19 or influenza outcomes. Of course, initial symptoms may be similar, e.g. fever, sore throat, muscle pain and cough, and in accordance with this we observed that >80% of patients were tested simultaneously (on the same date) for COVID-19 and influenza. These circumstances highlight the validity of our comparison of COVID-19 and influenza.

Although beyond the scope of our study, it is important to acknowledge the burden of infections not requiring hospitalization, as well as the burden of post-infection sequelae from COVID-19. The societal burden may include more frequent use of primary care services or consequences of absence from work.^41-43^

In conclusion, during the first two post-pandemic years COVID-19 represented a greater disease burden than influenza in Denmark, with more hospitalisations and deaths, and slightly more severe disease primarily among non-vaccinated and comorbid patients. These results underscore the need for continued attention, public health efforts and resources to mitigate the ongoing impact of COVID-19.

## Supporting information

none

## Data Availability

Aggregated admission and mortality data are available for download from dashboards at Statens Serum Institut (www.ssi.dk). De-identified individual-level data are available for research upon reasonable request to Statens Serum Institut and the Danish Health Data Authority and within the framework of the Danish data protection legislation and any required permission from relevant authorities. Applications should be submitted to Forskerservice (https://sundhedsdatastyrelsen.dk/da/forskerservice) where they will be reviewed on the basis of relevance and scientific merit. Data are available now, with no defined end date.

## Contributors

All authors contributed to study conception and design and interpretation of the data. IBS did the statistical analyses. PB and IBS had access to the underlying data, and drafted the manuscript. All authors provided critical revisions and final approval for the decision to submit for publication.

## Acknowledgments

This study was conducted as part of the Danish COVID-19 surveillance with governmental financial support.

