## Supplementary material for "The post-pandemic hospital and mortality burden of COVID-19 compared with influenza: A national cohort study in Denmark, May 2022 to June 2024": none

**Text A: Analyses of nationwide disease burden using incidence rate ratios of admissions of COVID-19 versus influenza**

All individuals living in Denmark during the study period were followed from May 16, 2022, until admission with COVID-19 or influenza, death, emigration, disappearance from national registers or the end of study period on June 7, 2024, whichever occurred first. Admissions and person time were aggregated, and negative binomial regression was employed to estimate adjusted incidence rate ratios (aIRRs) with 95% confidence intervals (CIs) to compare admission rates for COVID-19 and Influenza. Adjustments were made for sex and age group (0-39, 40-69, 70+). Interaction analyses were carried out, to assess whether the effect of disease varied by the following variables: sex, age group (0-39, 40-64 65+), year (2022/2023, 2023/2024) and season (summer and fall 2022, winter and spring 2022-2023, summer and fall 2023, winter and spring 2023-2024). Both main and interaction effects were included in these models. P-values for testing the homogeneity of effect were obtained from a likelihood ratio test comparing the goodness of fit between models with only main effect and models that included interaction terms. A test for overdispersion based on the simulated residuals was performed for all models.^44^

**Text B: Brief description of mass vaccination and circulating virus strains at the end of the pandemic**

During the pandemic between 2020 and until January 2022, the primary COVID-19 vaccines targeting the ancestral strain of SARS-CoV-2 had been administered to more than 80% adults aged ≥18 years in Denmark. In the winter season 2021-22, just before end of the pandemic, more than 60% of all adults received at least one COVID-19 booster dose, while to protect against influenza more than 80% of adults above 65 years of age received the quadrivalent influenza booster dose targeting the A(H1N1), A(H3N2), B-Victoria , and B-Yamagata strains.^20^ In February, 2022, when SARS-CoV-2 infections were decreasing with omicron being the dominant variant, a steep increase in influenza A infections were observed that peaked in March with the dominating subtype being A(H3N2) (98.9%) and only very few A(H1N1) (0.97%) and influenza B.^22^

**Figure S1:** Flowchart of study population.

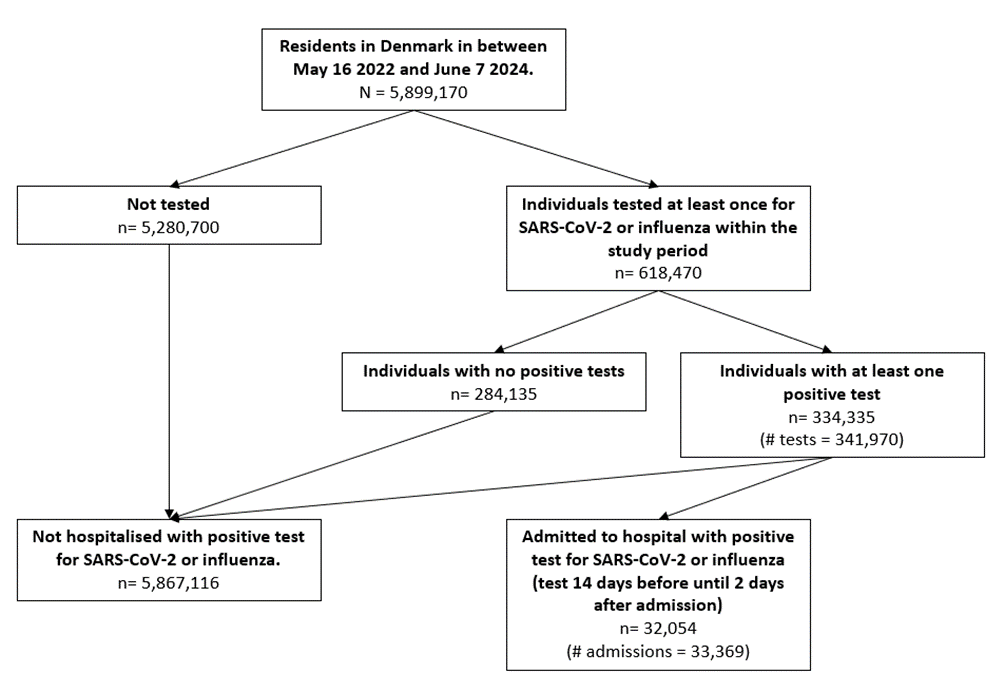

**Tabel S1:** Overview of variables used in the propensity score.

| **Variable** | **Categorization** | **Data source** |
| --- | --- | --- |
| Disease | ”COVID-19”, ”Influenza” | Danish Microbiology Database |
| Sex | ”Male”, ”Female” | Civil Registration System |
| Age at test | ”0-9”, ”10-19”, ”20-29”, ”30-39”, ”40-49”, ”50-59”, ”60-69”, ”70-79”, ”80-89”, ”90+” | Civil Registration System |
| Season of test | Spring, Winter | Danish Microbiology Database |
| Tilhørsforhold | “Danish”, “Born abroad”, “2. generation immigrant” | Civil Registration System |
| Region | Captial Region of Denmark, Zealand, Central Denmark, Northern Denmark, Southern Denmark | Civil Registration System |
| Long term care facility for elderly | “Yes”, “No | LTCF address database |
| Cardiovascular disease | “Yes”, “No | National Patient Registry |
| Respiratory disease | “Yes”, “No | National Patient Registry |
| Cancer | “Yes”, “No | National Patient Registry |
| Neurological disease | “Yes”, “No | National Patient Registry |
| Nephrological disease | “Yes”, “No | National Patient Registry |
| Hematological disease | “Yes”, “No | National Patient Registry |
| Immunological disease | “Yes”, “No | National Patient Registry |
| Asthma | “Yes”, “No | Register of Selected Chronic Diseases and Severe Mental Disorders |
| Dementia | “Yes”, “No | Register of Selected Chronic Diseases and Severe Mental Disorders |
| Diabetes, type 1 | “Yes”, “No | Register of Selected Chronic Diseases and Severe Mental Disorders |
| Diabetes, type 2 | “Yes”, “No | Register of Selected Chronic Diseases and Severe Mental Disorders |
| Chronic obstructive pulmonary disease | “Yes”, “No | Register of Selected Chronic Diseases and Severe Mental Disorders |
| Rheumatoid arthritis | “Yes”, “No | Register of Selected Chronic Diseases and Severe Mental Disorders |
| Osteoporosis | “Yes”, “No | Register of Selected Chronic Diseases and Severe Mental Disorders |
| Schizophrenia | “Yes”, “No | Register of Selected Chronic Diseases and Severe Mental Disorders |
| COVID-19 vaccination in the 180 days prior to test | “Yes”, “No | National Vaccination Registry |
| Influenza vaccination in the 180 days prior to test | “Yes”, “No | National Vaccination Registry |

**Table S2.** Overview of the PCR-testing patterns for COVID-19 and

influenza among hospitalized patients in the study.

|  | COVID-19 | Influenza |
| --- | --- | --- |
|  | n (%) | n (%) |
| **Tests performed** |  |  |
| Only for COVID-19 | 4413 (17.9%) | 0 (0.0%) |
| Only for influenza | 0 (0%) | 193 (2.2%) |
| Both* | 20274 (82.1%) | 8489 (97.8%) |

*Test for COVID-19 and influenza performed on the same day

**Tabel S3: Summary of length of hospital stay for patients with COVID-19 vs. influenza.**

|  | **Mean** |  | **Percentiles** | | | | |
| --- | --- | --- | --- | --- | --- | --- | --- |
|  |  |  | 5% | 25% | 50% | 75% | 95% |
| COVID-19 | 4.71 |  | 1 | 1 | 3 | 6 | 13 |
| Influenza | 4.08 |  | 1 | 1 | 3 | 5 | 12 |

**Table S4**. Characteristics and adjusted incidence rate ratios of admissions (n=33,369) and mortality (n=2,915) comparing patients with COVID-19 and influenza among the Danish population (n=5.9 mio., 11.9 million person-years of follow-up), May 2022 to June 2024.

|  | **Admission**  (positive virus PCR test up to 14 days before or 2 days after admission date) | | |  | **Mortality**  (30-day mortality after admission date) | |
| --- | --- | --- | --- | --- | --- | --- |
|  | COVID-19 | Influenza | SARS-CoV-2 vs. influenza: |  | COVID-19 | Influenza |
|  | n (%) | n (%) | Adjusted IRR* (95% CI) |  | n (%) | n (%) |
| **Total** | 24687 (100%) | 8682 (100%) | 2.01 (1.37, 2.95) | Total | 24687 (100%) | 8682 (100%) |
| **Year** |  |  |  |  |  |  |
| 2022/2023 | 16181 (65.54%) | 3973 (45.76%) | 2.63 (1.72, 4.01) |  | 1510 (63.1%) | 212 (40.61%) |
| 2023/2024 | 8506 (34.46%) | 4709 (54.24%) | 1.36 (0.90, 2.06) |  | 883 (36.9%) | 310 (59.39%) |
|  |  |  | P = 0.01 |  |  |  |
| **Season** |  |  |  |  |  |  |
| Summer-fall 2022 | 9210 (37.31%) | 173 (1.99%) | 38.67 (24.95, 59.96) |  | 796 (33.26%) | 10 (1.92%) |
| Winter-spring 2022/2023 | 7239 (29.32%) | 3820 (44%) | 1.17 (0.77, 1.76) |  | 736 (30.76%) | 202 (38.7%) |
| Summer-fall 2023 | 4271 (17.3%) | 213 (2.45%) | 14.23 (9.23, 21.94) |  | 376 (15.71%) | 13 (2.49%) |
| Winter-spring 2023/2024 | 3967 (16.07%) | 4476 (51.55%) | 0.69 (0.46, 1.03) |  | 485 (20.27%) | 297 (56.9%) |
|  |  |  | P < 0.001 |  |  |  |
| **Sex**** |  |  |  |  |  |  |
| Female | 11446 (46.36%) | 4406 (50.75%) | 1.87 (1.10, 3.18) |  | 1017 (42.5%) | 262 (50.19%) |
| Male | 13241 (53.64%) | 4276 (49.25%) | 2.16 (1.26, 3.69) |  | 1376 (57.5%) | 260 (49.81%) |
|  |  |  | P = 0.71 |  |  |  |
| **Age at study start** |  |  |  |  |  |  |
| 0-39 | 2053 (8.32%) | 2093 (24.11%) | 0.98 (0.75, 1.30) |  | 11 (0.46%) | 15 (2.87%) |
| 40-69 | 4362 (17.67%) | 2274 (26.19%) | 1.92 (1.46, 2.52) |  | 181 (7.56%) | 70 (13.41%) |
| 70+ | 18272 (74.01%) | 4315 (49.7%) | 4.30 (3.28, 5.63) |  | 2201 (91.98%) | 437 (83.72%) |
|  |  |  | P < 0.001 |  |  |  |

**IRR**, incidence rate ratio, e.g. the ratio of the national population incidence rates of SARS-CoV-2 versus influenza admissions.

*IRR adjusted for sex and age (0-39, 40-69, 70+).

**Due to low counts, IRRs are adjusted for sex and age (0-69, 70+).

**Table S5:** Detailed characteristics of admissions (n=33,369) of patients with COVID-19 or influenza, Denmark, May 16 2022 and June 27 2024.

|  | **Not influenza season** (i.e. summer season or June – November) | |  | **Influenza season** (i.e. winter season or December – May) | |
| --- | --- | --- | --- | --- | --- |
|  | **Influenza** | **COVID-19** |  | **Influenza** | **COVID-19** |
|  | n (%) | n (%) |  | n (%) | n (%) |
| **Total** | 377 (100%) | 13154 (100%) |  | 8305 (100%) | 11533 (100%) |
| **Sex** |  |  |  |  |  |
| Female | 192 (50.9%) | 6052 (46.0%) |  | 4216 (50.8%) | 5394 (46.8%) |
| Male | 185 (49.1%) | 7102 (54.0%) |  | 4089 (49.2%) | 6139 (53.2%) |
| **Age at test** |  |  |  |  |  |
| >40 | 88 (23.3%) | 1158 (8.8%) |  | 1939 (23.3%) | 831 (7.2%) |
| 40-65 | 96 (25.5%) | 2209 (16.8%) |  | 2056 (24.8%) | 1919 (16.6%) |
| >= 65 | 193 (51.2%) | 9787 (74.4%) |  | 4310 (51.9%) | 8783 (76.2%) |
| **Year** |  |  |  |  |  |
| 2022-2023 | 157 (41.6%) | 8858 (67.3%) |  | 3836 (46.2%) | 7591 (65.8%) |
| 2023-2024 | 220 (58.4%) | 4296 (32.7%) |  | 4469 (53.8%) | 3942 (34.2%) |
| **Region** |  |  |  |  |  |
| Capital Region of Denmark | 179 (47.5%) | 4229 (32.2%) |  | 2835 (34.1%) | 3740 (32.4%) |
| Central Denmark Region | 59 (15.6%) | 2362 (18.0%) |  | 1739 (20.9%) | 2244 (19.5%) |
| North Denmark Region | 35 (9.3%) | 1546 (11.8%) |  | 926 (11.2%) | 1125 (9.8%) |
| Region Zealand | 63 (16.7%) | 2265 (17.2%) |  | 1264 (15.2%) | 1963 (17.0%) |
| Region of Southern Denmark | 41 (10.9%) | ≤2747 (20.9%) |  | ≤1537 (18.5%) | ≤2456 (21.3%) |
| Missing | 0 (0%) | ≤5 (0.0%) |  | ≤5 (0.0%) | ≤5 (0.0%) |
| **COVID-19 vaccination** |  |  |  |  |  |
| Not vaccinated in 180 days prior to positive test | 289 (76.7%) | 10791 (82.0%) |  | 4446 (53.5%) | 5631 (48.8%) |
| Vaccinated in the 180 days prior to test | 88 (23.3%) | 2363 (18.0%) |  | 3859 (46.5%) | 5902 (51.2%) |
| **Influenza vaccination** |  |  |  |  |  |
| Not vaccinated in 180 days prior to positive test | 304 (80.6%) | 11356 (86.3%) |  | 4556 (54.9%) | 5588 (48.5%) |
| Vaccinated in the 180 days prior to test | 73 (19.4%) | 1798 (13.7%) |  | 3749 (45.1%) | 5945 (51.5%) |
| **Long term care facility for elderly** |  |  |  |  |  |
| No | 349 (92.6%) | 11332 (86.1%) |  | 7598 (91.5%) | 9708 (84.2%) |
| Yes | 28 (7.4%) | 1822 (13.9%) |  | 707 (8.5%) | 1825 (15.8%) |
| **Any comorbidity** |  |  |  |  |  |
| No | 110 (29.2%) | 3021 (23.0%) |  | 2448 (29.5%) | 2432 (21.1%) |
| Yes | 267 (70.8%) | 10133 (77.0%) |  | 5857 (70.5%) | 9101 (78.9%) |
| **Cardiac disease** |  |  |  |  |  |
| No | 248 (65.8%) | 7150 (54.4%) |  | 5565 (67.0%) | 6048 (52.4%) |
| Yes | 129 (34.2%) | 6004 (45.6%) |  | 2740 (33.0%) | 5485 (47.6%) |
| **Respiratory disorder** |  |  |  |  |  |
| No | 252 (66.8%) | 8527 (64.8%) |  | 5197 (62.6%) | 6797 (58.9%) |
| Yes | 125 (33.2%) | 4627 (35.2%) |  | 3108 (37.4%) | 4736 (41.1%) |
| **Cancer** |  |  |  |  |  |
| No | ≤372 (98.7%) | ≤12963 (98.5%) |  | 8218 (99.0%) | 11355 (98.5%) |
| Yes | ≤5 (1.3%) | ≤191 (1.5%) |  | 87 (1.0%) | 178 (1.5%) |
| **Neurological disease** |  |  |  |  |  |
| No | 359 (95.2%) | 11978 (91.1%) |  | 7742 (93.2%) | 10416 (90.3%) |
| Yes | 18 (4.8%) | 1176 (8.9%) |  | 563 (6.8%) | 1117 (9.7%) |
| **Nephrological disease** |  |  |  |  |  |
| No | 350 (92.8%) | 12070 (91.8%) |  | 7768 (93.5%) | 10490 (91.0%) |
| Yes | 27 (7.2%) | 1084 (8.2%) |  | 537 (6.5%) | 1043 (9.0%) |
| **Haematological disease** |  |  |  |  |  |
| No | 361 (95.8%) | 12334 (93.8%) |  | 7899 (95.1%) | 10669 (92.5%) |
| Yes | 16 (4.2%) | 820 (6.2%) |  | 406 (4.9%) | 864 (7.5%) |
| **Immunological disease** |  |  |  |  |  |
| No | 365 (96.8%) | 12710 (96.6%) |  | 8014 (96.5%) | 11085 (96.1%) |
| Yes | 12 (3.2%) | 444 (3.4%) |  | 291 (3.5%) | 448 (3.9%) |
| **Asthma** |  |  |  |  |  |
| No | 314 (83.3%) | 12267 (93.3%) |  | 7327 (88.2%) | 10908 (94.6%) |
| Yes | 63 (16.7%) | 887 (6.7%) |  | 978 (11.8%) | 625 (5.4%) |
| **Dementia** |  |  |  |  |  |
| No | 366 (97.1%) | 12592 (95.7%) |  | 8094 (97.5%) | 11154 (96.7%) |
| Yes | 11 (2.9%) | 562 (4.3%) |  | 211 (2.5%) | 379 (3.3%) |
| **Diabetes, type 1** |  |  |  |  |  |
| No | ≤372 (98.7%) | ≤13038 (99.1%) |  | 8182 (98.5%) | 11447 (99.3%) |
| Yes | ≤5 (1.3%) | ≤116 (0.9%) |  | 123 (1.5%) | 86 (0.7%) |
| **Diabetes, type 2** |  |  |  |  |  |
| No | 322 (85.4%) | 11141 (84.7%) |  | 7195 (86.6%) | 10045 (87.1%) |
| Yes | 55 (14.6%) | 2013 (15.3%) |  | 1110 (13.4%) | 1488 (12.9%) |
| **Chronic obstructive pulmonary disease** |  |  |  |  |  |
| No | 294 (78.0%) | 10853 (82.5%) |  | 6431 (77.4%) | 9471 (82.1%) |
| Yes | 83 (22.0%) | 2301 (17.5%) |  | 1874 (22.6%) | 2062 (17.9%) |
| **Rheumatoid arthritis** |  |  |  |  |  |
| No | 368 (97.6%) | 12696 (96.5%) |  | 8083 (97.3%) | 11202 (97.1%) |
| Yes | 9 (2.4%) | 458 (3.5%) |  | 222 (2.7%) | 331 (2.9%) |
| **Osteoporosis** |  |  |  |  |  |
| No | 323 (85.7%) | 11332 (86.1%) |  | 7340 (88.4%) | 10096 (87.5%) |
| Yes | 54 (14.3%) | 1822 (13.9%) |  | 965 (11.6%) | 1437 (12.5%) |
| **Schizophrenia** |  |  |  |  |  |
| No | ≤377 (99%) | ≤12996 (98.8%) |  | 8197 (98.7%) | 11414 (99.0%) |
| Yes | ≤5 (1%) | ≤158 (1.2%) |  | 108 (1.3%) | 119 (1.0%) |
